# Evidence on the role of journal editors in the COVID19 infodemic – metascientific study analyzing COVID19 publication rates and patterns

**DOI:** 10.1101/2022.01.23.22269716

**Authors:** Gabriel de Araújo Grisi, João de Deus Barreto Segundo, Camila Verônica Souza Freire, Denise Silva Matias, Mariana Correia Moreira Cruz, Larrie Rabelo Laporte, Daniel Oliveira Medina da Silva, Thiago Masashi Taniguchi, Letícia Escorse Requião, Bruno Teixeira Goes, Luis Claudio Lemos Correia

**Affiliations:** Center for Evidence-based Medicine at Escola Bahiana de Medicina e Saúde Pública (Salvador, Brazil)

**Author notes:** **Correspondence and requests** should be addressed to Luis Claudio Lemos Correia at or at the Center for Evidence-Based Medicine, Escola Bahiana de Medicina e Saúde Pública (Dom João VI Avenue, 275, Brotas. Salvador, Bahia, Brazil. Postal code 40290-000).

**Keywords:** Scientific communication, Scientific publications: periodicals, Scientific publications: preprints, COVID19, Influenza

## Abstract

**Objective:** Infodemic, a neologism characterizing an excess of fast-tracked low quality publications, has been employed to depict the scientific research response to the COVID19 crisis. The concept relies on the presumed exponential growth of research output. This study aimed to test the COVID19 infodemic claim by assessing publication rates and patterns of COVID19-related research and a control, a year prior.

**Design:** A Reproduction Number of Publications (R_p_) was conceived. It was conceptualized as a division of a week incidence of publications by the average of publications of the previous week. The publication growth rates of preprint and MEDLINE-indexed peer-reviewed literature on COVID19 were compared using the correspondent Influenza output, a year prior, as control. R_p_ for COVID19 and Influenza papers and preprints were generated and compared and then analyzed in light of the respective growth patterns of their papers and preprints.

**Main outcomes:** Output growth rates and Reproduction Number of Publications (R_p_).

**Results:** COVID19 peer-reviewed papers showed a fourteen fold increase compared to Influenza papers. COVID19 papers and preprints displayed an exponential growth curve until the 20th week. COVID19 papers displayed R_p_=3.17±0.72, while the control group presented R_p_=0.97±0.12. Their preprints exhibited R_p_=2.18±0.54 and R_p_=0.97±0.27 respectively, with no evidence of exponential growth in the control group, as its R_p_ remained approximately one.

**Conclusions:** COVID19 publications displayed an epidemic pattern. As the growth patterns of COVID19 peer-reviewed articles and preprints were similar, and the majority of the COVID19 output came from indexed journals, not only authors but also editors appear to had played a significant part on the infodemic.

**Review protocol:** https://osf.io/q3zkw/?view_only=ff540dc4630b4c6e9a2639d732047324

**Ethical aspects:** No ethical clarence was required as all analyzed data were publicly available.

**SUMMARY BOX**
**1. What is already known about this subject?**
Much has been commented on 2020’s excess of publications on COVID19. Independent studies found evidence of increased volume and speed of publication, decreased methodological quality, and qualitative variations in peer review of COVID19 papers, when compared to the scholarly output from before the pandemic. This phenomenon has been branded an infodemic, a neologism implying an epidemic of low-quality information on COVID19 when high quality scientific reports to inform health policies would have been needed the most.
**2. What are the new findings?**
No study pushed the infodemic metaphor forward to analyze not only volume of publication but also publication rates comparing them to a control group as to clearly pinpoint an exponential phase of contagion in the infodemic (as it would take place in a real epidemic) through a mathematical analysis of the growth patterns and rates of those publications. In this paper, we were able to demonstrate that there has been an infodemic indeed and that the editor population was as susceptible to the infodemic bug as the author population because the exponential phase was shaped not only by authors but mainly by editors from PubMed-indexed journals.
**3.How might it impact clinical practice in the foreseeable future?**
These results and conclusions are consequential to subsequent studies on rigor and depth of post publication peer review and on editorial practices within the life and health sciences research community.

## INTRODUCTION

The global scientific output has been doubling every nine years^1^ leaving the scientific community scrambling to keep up-to-date and to separate wheat from chaff^2-4^. Excessive publication is an long-established scientific community complaint, an issue the scientific journal was created to overcome by organizing and validating research^5-8^. Notwithstanding, quality complaints encompassed in the concept of avoidable research waste is a recent issue. Awareness regarding this topic has been increasing for the past thirty years^9-11^, giving rise to the notion that perhaps the incentives of the scientific ecosystem could have been skewing publication output towards novelty and quantity and away from reliability and quality^2,3,11^. If pressures for publishing innovation were to be considered indeed a strong drive behind the current scientific output and the average methodological quality of biomedical research had already been deemed uncommendable before the pandemic^2-4,9-11^, what would happen should the scholarly publishing ecosystem find itself under even more pressure caused by a worldwide sanitary crisis?

Mounting evidence from independent studies have been suggesting that the publication ecosystem appears to have been rushing the publication of lower quality COVID19-related papers in journals in comparison to pre-pandemic times: the time elapsed from article submission to publication of COVID19-related articles in journals has accelerated^12-17^ while there is evidence that the overall methodological quality of studies has decreased^15,18^. The median time to reach final acceptance stage was found to be eight times faster for COVID-related peer-reviewed articles in comparison to papers on other issues by independent studies^15,16,19^. In a sample of PubMed-indexed journals, 10% of COVID-related studies were found to have been accepted within two days of submission^16^. A study on open peer review from two high tier journals found qualitative distinctions in COVID19-related papers when compared to non-related ones. The prior category usually underwent a single round of peer review, displayed less propensity to request further data and more propensity to request authors to tone down claims and conclusions. Before the pandemic, they usually underwent two or more rounds of review^19^.

It has also been found that, in 2020, 27% of the active authors from the SCOPUS database published COVID19 research in a subfield discipline that was not among the top three subfield disciplines in which they had published most commonly during their career. Approximately one in seven active scientists publishing in English-speaking high and middle tier journals rapidly adjusted their portfolio to procure publications on COVID19. Those authors were found across twenty one major fields of SCOPUS and included experts specializing in fishery, ornithology, entomology and architecture that had published on COVID19 in 2020^20^. Furthermore, COVID19 manuscripts were uploaded as preprints concurrently to their submission to journals, implying their authors generally did not specifically pursue the pre-submission feedback when posting their research as preprint^21^. It is noteworthy, however, there is no evidence COVID19 preprint authors published outside of their expertise^22^. Those findings point to an informational phenomenon that could maybe be contagious as other information phenomena have proved to be. A famous example is the Matthew Effect, in which citations are associated with more citations over time^23,24^, implying citations attract citations, as attention attracts attention. The ongoing publishing phenomenon has been referred to by the scholarly community as an infodemic^12,25^. But, could the deluge of papers from journals and preprints indeed be framed as an infodemic in which COVID19 publications would have lead to even more COVID19 publications exponentially? As compared to what control-group and to which period of time? And, if so, what could have been the participation of journals and preprint servers in the phenomenon? Have the COVID19-related output rates been following some sort of epidemic pattern or was the term infodemic just a catching metaphor? Were preprints to blame? And if so, to what extent?

In view of this rationale, this study intended to evaluate whether COVID19-related publications followed an epidemic pattern, displaying expontential growth and a Reproduction Number of Publications (R_p_) above one, being the R_p_ defined as a division of a weekly incidence of publications by the average of publications of previous weeks. Additionally, it aimed to assess the behavior of preprint publication rates compared to their journal counterparts. To the best of our knowledge no such comparison has yet been made. Preprints have proved to be a relevant part of the scientific publication ecosystem, informing policy and being increasingly discussed in the lay press^22,26-28^. The authors understand the seriousness of the current crisis and their objective was to compare the publication ecosystem under normal circumstances to it under current pandemic pressures and, if possible, critically characterize and discuss an eventual infodemic through a thought experiment. Thus, publications on Influenza were chosen as the control group to test the infodemic claim because both Influenza and COVID19, albeit very different when it comes to severity and morbidity, are ongoing pandemics. Publications on MERS-CoV, Ebola virus disease or Zika virus were ruled unfit for comparison because their respective outbreaks were self-limited in time and to specific regions of the world thus unable to exert continuous pressure on the publication ecosystem as a whole.

## RESULTS

### Incidence of Publications

From January 2020, month of the first COVID19 publication, to February 6th 2021, the COVID19 output increased weekly to a total of 97,781 documents, a fourteen-fold increase compared to the Influenza output, which accumulated 6,936 documents for the equivalent period. COVID19 journal articles incidence increased until the 20^th^ week of 2020, weekly, from January to June, while Influenza output had remained stable over time a year prior (Figure 1). From the 20^th^ week to 52^nd^ week, the number of COVID19 journal articles became steady at 1,820 (SD ± 233.8). (Figures 1 and 2 in Appendix).

**Figure 1.**
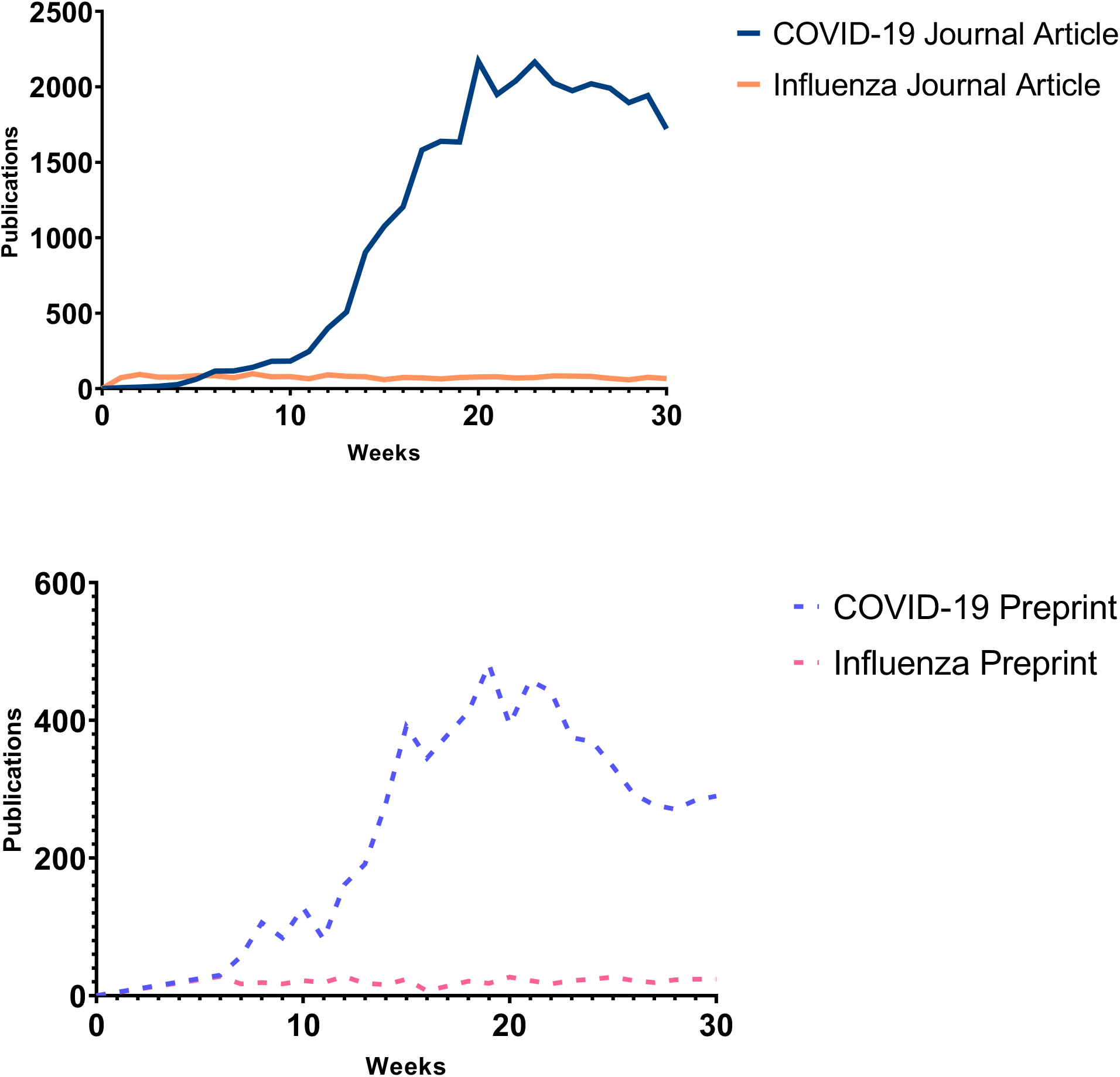
and 1a. COVID-19 and Influenza publications incidence per week.

**Figure 2.**
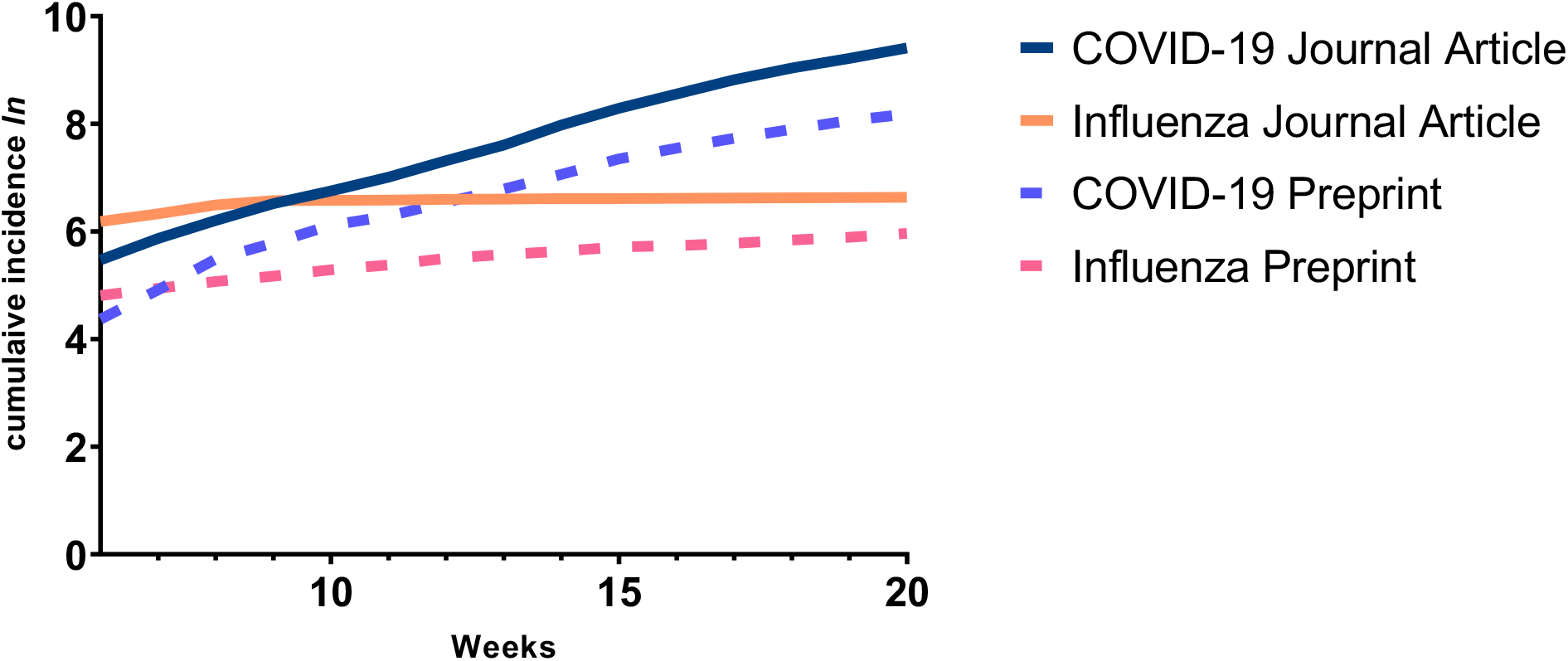
Log-transformed cumulative publications per week.

From the 1^st^ to the 30^th^ week in the 2020 timeframe, COVID19 preprints accounted for 6,960 documents and Influenza for 614 (Figure 1a). Incidence of COVID19 preprints also increased significantly until the 19^th^ week followed then by a fluctuating incidence number with downward trend. The control group on the other hand displayed stability in all analyzed time frames.

### Modeling Function of Publications over Time

For both COVID19 journal articles and preprints, the log transformation of the weekly cumulative number of publications displayed an exponential behavior from weeks 6 to 20 (Figure 2). The linear regressions described the data at this time-lapse with an R^2^=99% for journal articles and an R^2^=96% for preprints, through the weeks (t) with: *f*(*t*) = 0.28*t* + 3.9 and *f*(*t*) = 0.26*x* + *3*.*3* respectively.

Exponential coefficients (linear regression slope) of 0.28 (95% CI, 0.28-0.3) and 0.26 (95% CI, 0.23-0.29) for COVID19 journal articles and preprints were found respectively. The exponential models for publications (p) by time (t) were *p*(*t*) = 206.29*e*^0.284(*t*)^ for COVID19 journal articles and *p*(*t*) = 100.32*e*^0.258*t*^ for COVID19 preprints.

The Influenza control group displayed a stable linear growth pattern. At the cumulative semi-log function, Influenza journal articles and preprints presented a slope of 0.08 (95% CI, 0.07 – 0.09), R^2^=96%). Accordingly, if we analyze the cumulative incidence function over time (without any log transformation), a linear pattern with a slope of 75.8 (95% CI, 74 - 78) for journal articles and 18.8 (95% CI, 18-20) for preprints was found, each with an R^2^=99.8%. Consequently, the best function to represent the control group was not an exponential one.

### Publication Reproduction Analysis

The R_p_ for journal articles from the 6^th^ to the 20^th^ week showed statistically significant difference between COVID19 (3.14±0.71) and Influenza (0.96±0.12), p<0.01. Difference was also found for preprints: COVID19 (2.78±0.53) and Influenza (0.99±0.27), p<0.01. Within COVID19 and Influenza groups, R_p_ of preprints and journal articles did not differ: p=0.13 for COVID publications and p=0.64 for Influenza (Figure 3).

**Figure 3.**
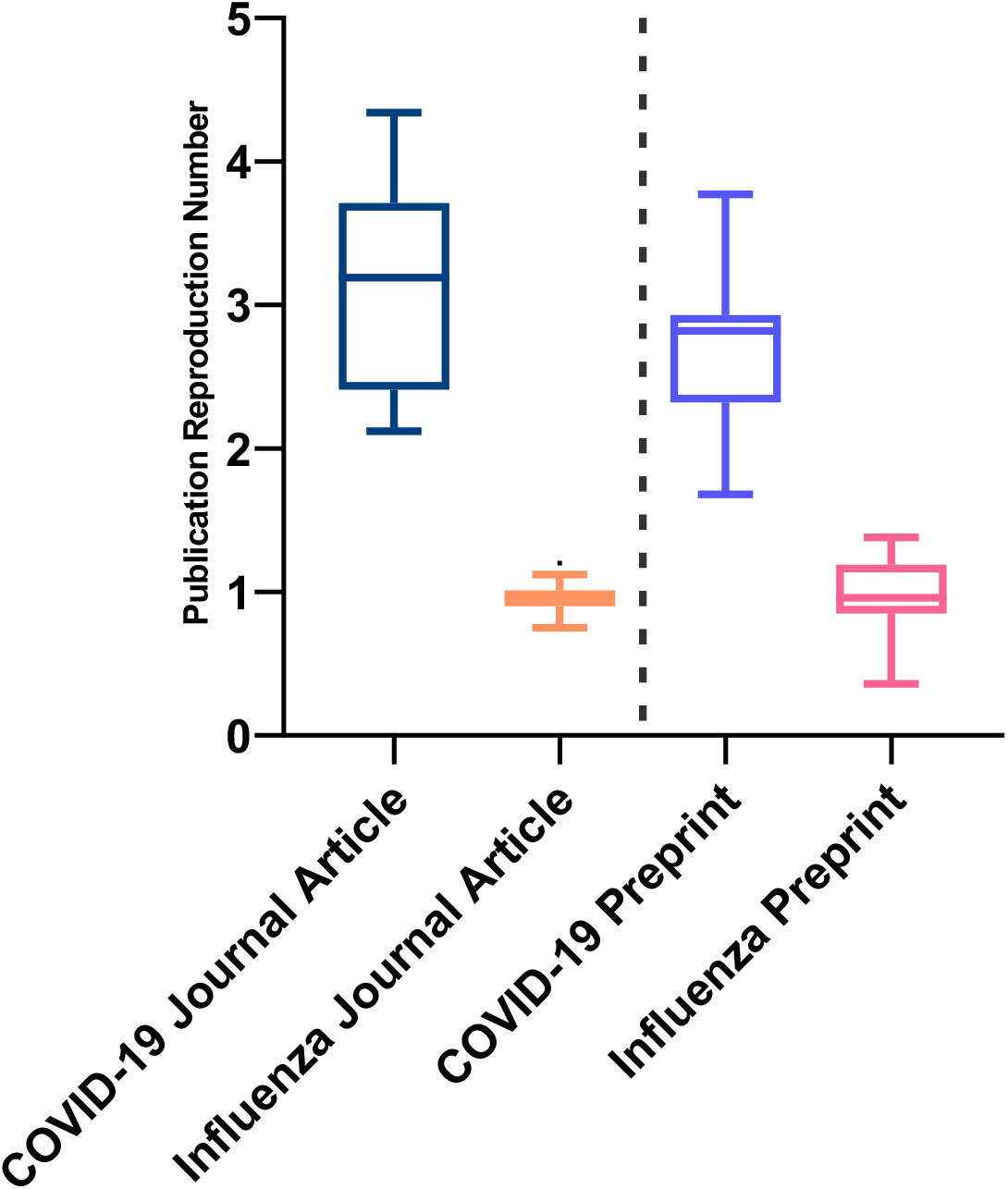
Boxplot showing R_p_ behavior for weeks 6 to 20.

Regarding R_p_ for journal articles and preprints, within a putative model where each article has perpetual influence over others, a result greater than one represented transmission as if an average of 3.14 secondary journal articles and 2.78 secondary preprints had been spawned from their predecessors. Comparatively, the Influenza control group displayed no transmission patterns, since its R_p_ was found to be approximately one (Figure 4). After the 20^th^ week, R_p_ for COVID19 journal articles leaned towards one, ending the exponential phase of the publication output growth (Figure 3 in the Appendix).

**Figure 4.**
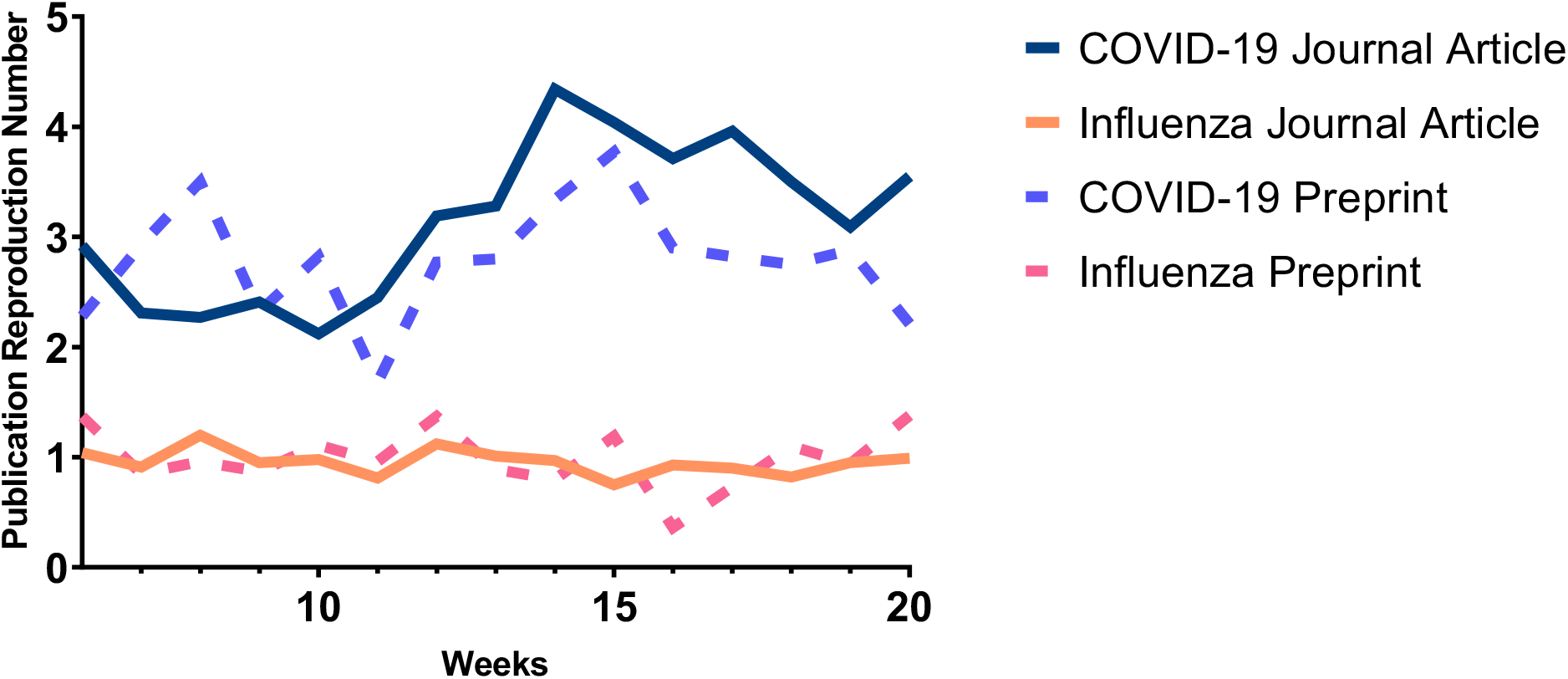
COVID-19 and Influenza R_p_ behavior per week until the 20^th^ week.

## DISCUSSION

Epidemics may result from a combination of transmissibility of an etiological agent, environmental context and susceptibility of a population. Our findings suggest that the deluge of COVID19-related papers and preprints could indeed be framed as an infodemic as compared to the control group, resulting from (a) a very transmissible bug, (b) exposed and susceptible individuals in (c) an environment that enhanced transmission due to the natural urgency to find solutions for the crisis and, possibly, pressure for publication and recognition. As the present findings show, it becomes likely that, in the early months of the pandemic, the more papers were published, the more authors fed into the cycle of transmission by publishing yet more papers both in traditional journals and in the preprint servers, exposing themselves and others.

It is interesting to note, however, that both preprint publication and journal publication displayed equivalent growth patterns suggesting that the publishing venues may have reacted to community pressures. In other words, as Influenza-related articles had been published in journals, they were published in preprint servers in a proportional frequency, displayed by the two superimposed growth curves with analogous patterns (Figure 4). As COVID19-related articles were published in journals so they were published in preprint servers in the same fashion, although following a steeper growth curve on both venues.

In view of that, we would do very well to keep in mind that, as a contaminated doorknob is no more responsible in transmission than the unwashed hand that touched the knob and then the face, the publication venues appear to have behaved as the scientific community shaped them to. However, because journals have placed themselves in the past two hundred years as official science adjudicators, and, according to our findings they were responsible for the bulk in COVID19 publication, they could be interpreted as being the main culprit behind the infodemic. Thus, it would appear that the infodemic bug contaminated not only authors, but also editors.

It could be speculated perhaps that journal editors may have decided to accelerate the publication process, publish more and, perhaps, expect the community to judge later, placing a lot of pressure on post-publication peer review, which may or may not have happened and is a topic for further investigation in other studies, being outside the scope of this thought experiment. Mainstream life and medical science preprint servers, on the other hand, played but a very small part in the infodemic while journals acted as the main superspreading venues as they are the main infrastructure that to this date dominates the scientific publication ecosystem.

On the other hand, a greater volume of journal articles does not necessarily imply they received greater attention, both in lay media and in scientific circles as to, by themselves, account for disinformation, misinformation and all in between in the COVID19 era. It is likewise not possible, within the scope of this experiment, to argue that preprints did not account for more disinformation than their journal counterparts. To the best of our knowledge, there is no strong empirical evidence swaying the argument either way. Any future analysis should consider the attention received by published papers and preprints in the form of citation, social media mentions and lay media coverage. A future infodemic investigation would do well to assess and compare Altmetrics and citations for journal articles and preprints employing also a control group. Considering preprints are readily accessible, it is plausible that they could have played an important part on the spread of the infodemic bug by means of social media and lay media coverage. However the debate is far from settled and in need of further evidence.

At this point, everyone is well aware that more publication does not equal better, more relevant and useful publication. The lower quality of journal articles during the pandemic has been identified by independent studies and, due to that, the quality of peer review is to be presumed lacking. Therefore the infodemic may as well have brought about a secondary wave of hazards such as cognitive fatigue and the possibility of having good, useful and relevant COVID19 research buried under a deluge of low quality research reports. It is not to be implied that all COVID19-era research has resulted from a desire for recognition in spite of quality but our findings, along with previous findings on the quality of pandemic publication output, make a strong case that recognition-seeking played a very strong part, adding strength to claiming the infodemic as an actual contagious informational (and, thus, behavioral) phenomenon among authors, not as a metaphor alone.

The infodemic bug is still active, there is no immunization against publishing urges and pressures yet. It is just not exponential anymore as it was in the early months of 2020, which is a good sign. This has not been the first time the scientific community has been at odds with the amount of research output it spawned. The journals themselves have been established so that scientists could cope with the amount of literature they had to keep up with in order to remain up-to-date back in the XVII century. Thus, one could argue that the cognitive overload after the bibliographic explosion exerted an adaptive pressure on the scientific publication ecosystem towards professionalization. The novelty in the COVID19 infodemic is the widespread use of preprint servers, which may play out as another adaptive pressure on the scientific publication ecosystem towards more openness and post-publication peer review. As the evidence showed, if journals published so much more and while presenting a significantly worse quality than in pre-pandemic times, perhaps more skepticism towards pre-publication peer review would be in order as well.

In light of the present findings, it seems the publication venues will be what authors and editors shape them to be, even more so than reviewers. Thus, there is no point in placing the blame onto structures and technologies and not tackling the real issue: pressure for publication and recognition in order to achieve career advancement, at the expense of methodological rigor and quality. This is the underlying complication the debate around research waste has been pointing out for the past thirty years. So, as we hope for our thought experiment to have shown, for both journals and preprint servers to provide fruitful high-quality public scientific debate by means of reliable research, both authors and editors need to put quality and transparency before speed and novelty and actively resist the infodemic bug themselves.

## METHODS

### Query

For articles from PubMed-indexed journals, Entrez Direct (EDirect) NIH/NLM application was employed for retrieving publication metadata from MEDLINE by E-utilities API^29,30^. MEDLINE metadata retrieval included papers from December 1^st^, 2019 to February 6^th^, 2021 for COVID19. The same interval one year prior was considered for Influenza papers because publications on that topic were presumed to have been suppressed in 2020 due to the current focus of the experts on the ongoing COVID19 pandemic. Thus, this study aimed for a more conservative approach as to not overestimate journal article output findings.

Preprint metadata was retrieved from the bioRxiv and medRxiv servers^31^, presumed to be the most prominent among the life sciences servers to this date^21,22,28^. The rationale was: if PubMed was to be considered a collection of high quality biomedical output, the equivalent collection among preprint servers would be both bioRxiv and medRxiv. The native advanced search tools connecting medRxiv and bioRxiv databases was employed for querying preprints^31,32^. Preprint metadata retrieval interval for COVID19 ranged from January 13^th^, 2020 (first occurrence) to July 28, 2020. The same interval, one-year prior, was considered for Influenza preprints.

To rule out the possibility of missing metadata in the EDirect output files, COVID19 EDirect queue results were compared to the PubMed queue results^33^. Comparison revealed no loss of articles from automatic extraction employing EDirect. Thus, to reduce the possibility of manual metadata collection inducing bias or loss of metadata, it was decided upon using the EDirect software package. Search strategies, data extraction and technical software aspects are detailed in the Appendix.

### Eligibility

All journal articles from EDirect queries were included. Preprints that came in mixed up within those search results and those articles that did not come with a full date of publication upon extraction were excluded. All bioRxiv and medRxiv search results for preprints were included as all of them have publication date. No metadata curation was performed at this point for any group. A simulation was run, excluding letters and opinion pieces from the published papers output, amounting to no expressive change in quantity of published pieces.

### Extraction and internal validity assessment

EDirect metadata from COVID19 and Influenza journal articles was downloaded into a Microsoft Excel readable document (Microsoft Corporation, United States of America). Influenza preprint metadata was exported into an Extensible Markup Language file from the preprint search result pages. COVID19 preprints were collected from the COVID19 dedicated page connecting the servers’ databases, accessed in July 28^th^, 2020^31^.

To assess the validity of employing a non-curated and automatic method of metadata extraction, the search strategy for journal articles was repeated at different timepoints, in which the PubMed output was compared to NCBI LitCovid output^34^. Due to that assessment, PubMed was confirmed to have 1.17 more publications on COVID19 than LitCovid did then, indicating a possible delay in the LitCovid update timeframe. Thus, it was decided PubMed was more reliable for an accurate coverage. Influenza metadata results were deemed not needed to be validated or curated due to the output being eight times smaller than its COVID19 counterpart.

Attribute variable *PubmedPubDate@pubmed*, a date that reflects inclusion in PubMed database, and *PubDate*, a date that reflects the issue date of publication, were extracted from PubMeb as quality control of *ArticleDate*, a variable which corresponds to the date the journal publisher has made an electronic version of the article available online. *PubmedPubDate@pubmed* PubDate was perceived to display loss in accuracy, with papers that seemed to have been added into the NLM collection, in bulk, days or weeks after their first online publication date in their journal websites. Thus, *ArticleDate* was chosen as variable of interest because it showed to be equivalent or slightly more nuanced than *PubmedPubDate@pubmed* and to *PubDate*, presenting itself as the most accurate choice^35^ (see Figure 4 in Appendix).

To enable the creation of a timeframe of incidence of publication, weeks were numbered automatically employing Excel, placing week 1 in January for all groups.

### Function appraisal

As every exponential function becomes linear when it is log-transformed, to allow for the testing of the hypothesis of exponential growth, a log transformation on the cumulative incidence variable was performed. Hence, the data linearity was appraised by linear regression throught its function slope, or growth rate (r), which is also present at the exponential function model *x*(*t*) = *ae*^*r*(*t*)^, and the determination coefficient (R^2^). Exponential model function was described using trendline equation in Excel. The growth pattern of the curve was analyzed and the steeper the inclination, the more pronounced the growth was, which characterized the multiplier effect (r) of the time independent variable as the power of the exponential function. Time intervals for analysis were later defined after evaluation of curve behavior and best function fit.

### Publication Reproduction Number

The well-known effective reproduction number (R_t_) helps project the epidemic growth patterns because it is an indicator of the contagiousness of an etiologic agent within a specific timeframe and environmental context. It has been established as the number of secondary cases a single case at a given time would generate^36^. Based that rationale, a parameter conceptually inspired by the effective reproduction number was conceived for this thought experiment: the Publication Reproducibility Number (R_p_). This parameter consisted in the division of the weekly incidence of publications by the average of previous weekly cases, where *i* is the first considered week for a given time-lapse and *n* is the total number of weeks within the analyzed timeframe, in which *s* stands for week incidence, and *c* is the current week incidence.

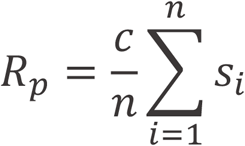

In the context of an infodemic, new cases were to be understood as additional published papers or preprints. This concept was conceived after the assumption that the desire to publish COVID19-related studies could have been amplified by the amount of COVID19 literature already published and the attention it thus received, which is supported by mounting independent evidence on author publication behavior in the COVID19-era^20,22^ and on the faulty reward system of scientific publication^2-4, 9, 10^.

As the infodemic metaphor went, to better define an epidemic of published papers and preprints, the *R*_*p*_ would paint a clearer picture where simple comparison of quantity of published papers and preprint output could not. So, within a putative model where each article had perpetual influence over the subsequent article generation, authors would influence -or, metaphorically, contaminate - other authors with a misplaced urge to spawn additional COVID19 papers. So, as this rationale went, the way the *R*_*p*_ had grown in the early months of the COVID19 pandemic could inform how much the average incidence of new weekly publications would be increasing or decreasing, supporting or refuting the infodemic claim.

### Data Analysis

Journal articles and preprints were compared by theme of publication (COVID19 or Influenza) and by type of publication (preprints or journal articles). The numerical variables were described as mean and standard deviation, after the data distribution appraisal. To compare the COVID19 and Influenza R_p_ means, the bilateral independent Student’s t-test was employed. Exponencial function growth rate was achieved throught the linear regression and described with confidence interval. For all analysis, p-value of less than 0.05 was taken as statistically significant. Statistical analysis and curve estimations were executed in SPSS Statistics version 14 (International Business Machines Corporation, United States of America), graphics were generated using GraphPad Prism 8.0.

## Data Availability

The datasets generated during and/or analysed during the current study are available from the corresponding author upon request.

## Code Availability

The codes generated and/or analysed during the current study are available in the Appendix.

## Funding

The authors received no funding whatsoever to conduct this study.

**Supplementary information is available for this paper and was inserted at the Appendix**.

## APPENDIX

**Figure A1.**
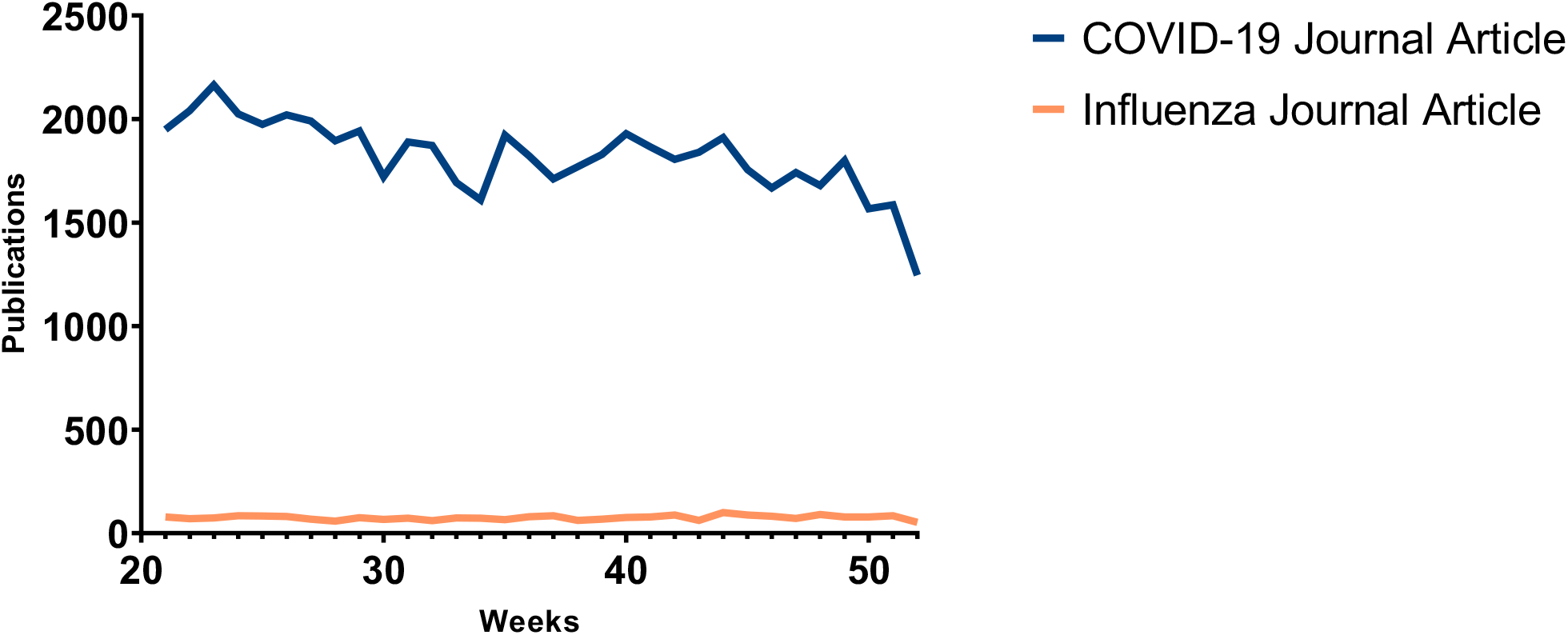
Incidence of articles from weeks 21 to 52.

**Figure A2.**
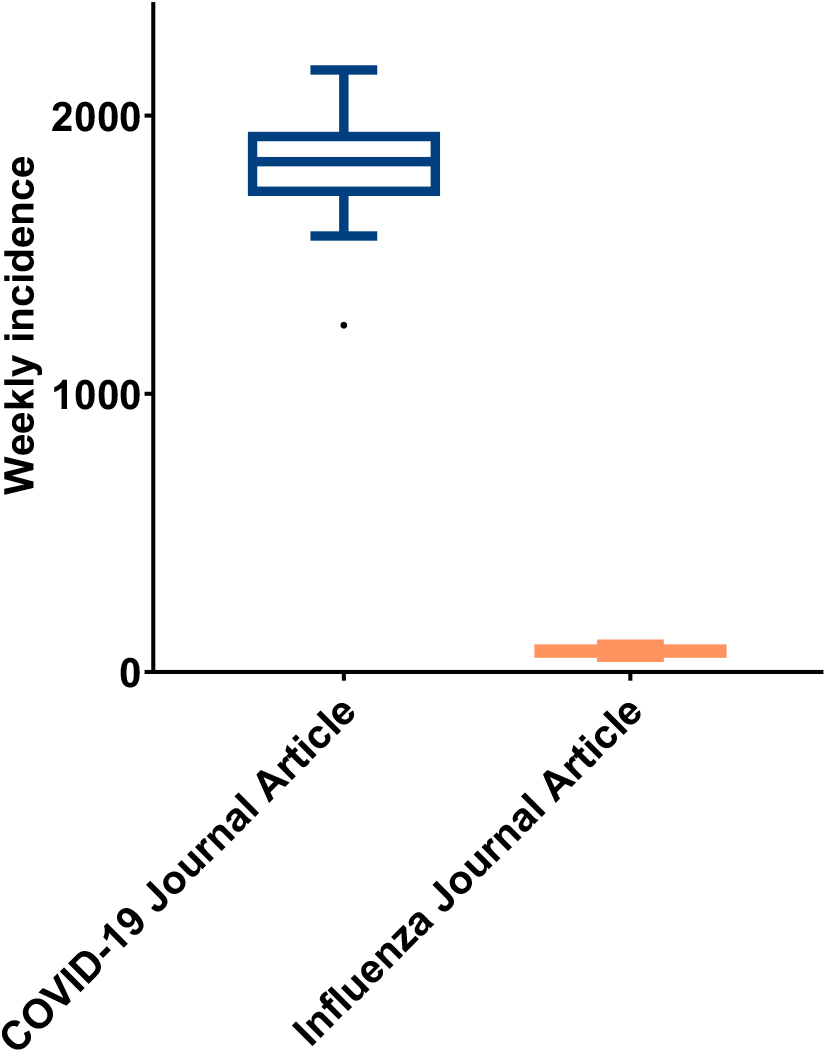
Incidence output of journal articles from weeks 21 to 52;

**Figure A3.**
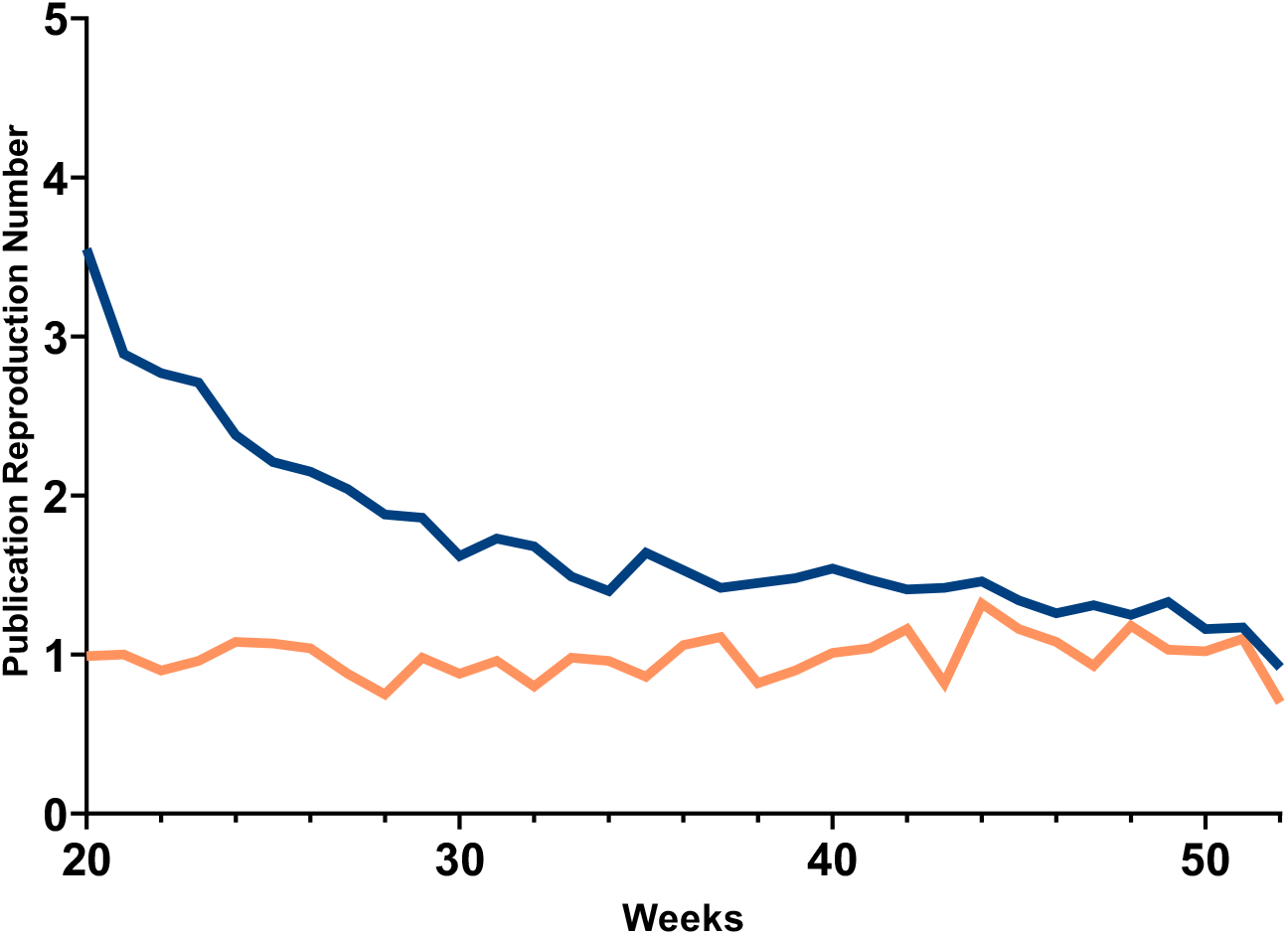
Journal articles R_p_ from weeks 20 to 52.

**Figure A4.**
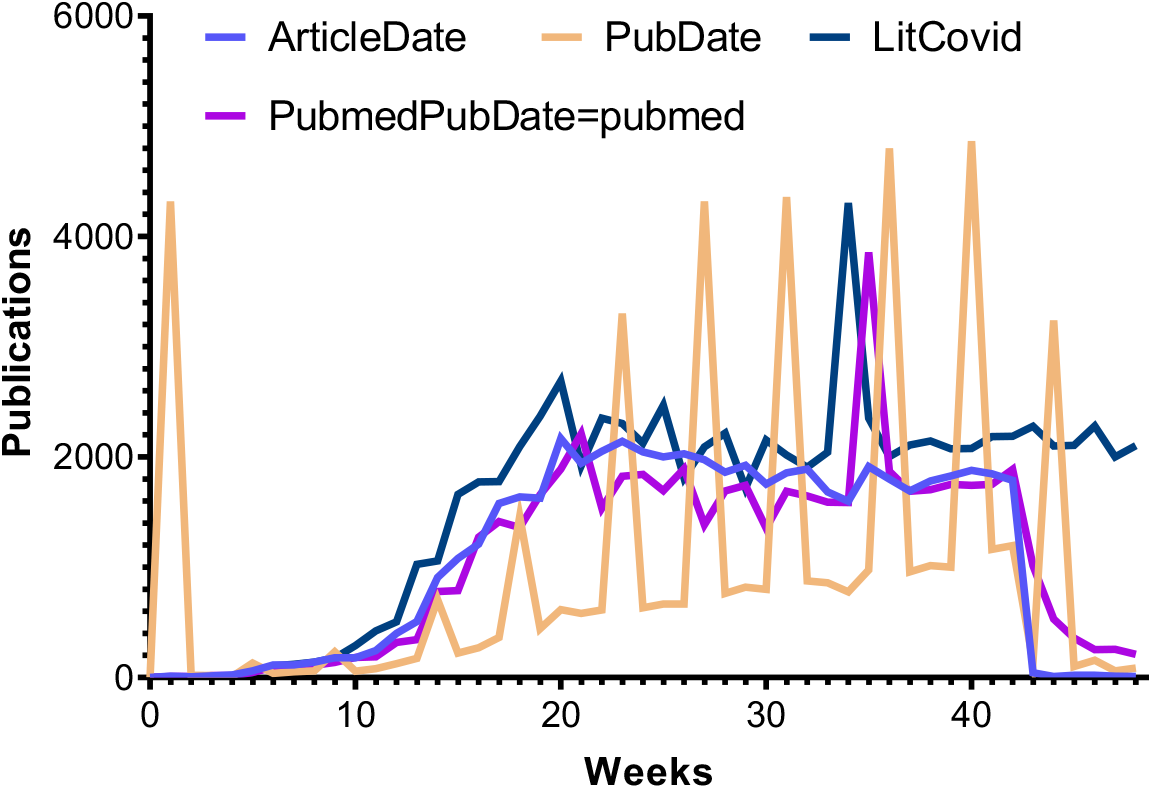
PubMed metadata plotted in weeks for the selection of a variable that would best describe the timeframe of metadata inclusion.

### Search strategy

- SARS-Cov-2 query

*((wuhan[All Fields] AND (“coronavirus”[MeSH Terms] OR “coronavirus”[All Fields])) AND 2019/12[PDAT]*: *2030[PDAT]) OR 2019-nCoV[All Fields] OR 2019nCoV[All Fields] OR COVID-19[All Fields] OR SARS-CoV-2[All Fields]* (1)

- Influenza query

*(“Human Influenzas” OR “Influenzas, Human” OR “Influenza” OR “Influenzas” OR “Human Flu” OR “Flu, Human” OR “Human Influenza” OR “Influenza in Humans” OR “Influenza in Human” OR “Grippe”) AND (2018/12[PDAT]*: /last retrieval month in 2019*\[PDAT])*

### Data Retrieval, extraction, and formatting

#### Journal Articles

E-Utilities in EDirect(2) UNIX terminal environment in Cygwin(3) software were used at the tools version as follows from installed confirmation message: esearch version: 13.95, xtract version: 14.6.

### Code string

#### SARS-Cov-2

$ esearch -db pubmed -query “((wuhan[All Fields] AND (“coronavirus”[MeSH Terms] OR “coronavirus”[All Fields])) AND 2019/12[PDAT]: 2030[PDAT]) OR 2019-nCoV[All Fields] OR 2019nCoV[All Fields] OR COVID-19[All Fields] OR SARS-CoV-2[All Fields]” | efetch -db pubmed -format xml | xtract -pattern PubmedArticle -sep “/” -def “N/A” -element MedlineCitation/PMID ArticleTitle ISOAbbreviation PublicationType Language Country ArticleDate/Year,ArticleDate/Month,ArticleDate/Day PubDate/Year,PubDate/Month,PubDate/Day -block PubMedPubDate -if PubMedPubDate@PubStatus -equals pubmed -sep “/” -def “N/A” -element PubMedPubDate/Year,PubMedPubDate/Month,PubMedPubDate/Day -block ArticleId -if ArticleId@IdType -equals doi -element ArticleId > sars2_database.txt

### Influenza

$ epost -input *(downloaded PMID list of previous PubMed search)*.txt -db pubmed | efetch -db pubmed -format xml | xtract -pattern PubmedArticle -sep “/” -def “N/A” -element MedlineCitation/PMID ArticleTitle ISOAbbreviation PublicationType Language Country ArticleDate/Year,ArticleDate/Month,ArticleDate/Day PubDate/Year,PubDate/Month,PubDate/Day -block PubMedPubDate -if PubMedPubDate@PubStatus -equals pubmed -sep “/” -def “N/A” -element PubMedPubDate/Year,PubMedPubDate/Month,PubMedPubDate/Day -block ArticleId -if ArticleId@IdType -equals doi -element ArticleId > iflz_database.txt

### Enhancing visualization and data processing

- *Microsoft Excel* imported through *Power Query* the data from *EDirect* text format articles outputs or downloaded *xml* preprints archives.
- Duplicates and references with *PublicationsType* variable equal to “Preprint” were automatically excluded.
- Several formulas were used to extract and plot the number of publications per week, as follows:

*Reference example cell containing date information: Gx, where x is a row*.

K = Standard Month (a list for formula reference);

L = Standard Week (a list for formula reference);

M = Article Date (the excel reference data extracted from the original fetched data); =VALUE(sars2_database[@ArticleDate])

N = Year of the M value; =YEAR(Mx)

O = Week of the M value; =ISOWEEKNUM(Mx)

P = Countable Week (filtered weeks, excluding results out of interest range – 2019 to 2021 – also where 2021 weeks are added 52 – total number of 2020 weeks – to continue the number of weeks as the first being the 1^st^ of January 2019 week, and the last is the current week in 2021; =IFS(YEAR(Mx)=2020,Ox,YEAR(Mx)=2021,52+Ox,AND(YEAR(Mx)=2019,Ox<53),9999, YEAR(Mx)<=2018,9999)

Q = PubWeek (the incidence number in the referred week); =COUNTIF(P:P,Lx)

R = Cumulative value of incidence per week; =(SUM(Q2:Qx))*

S = Neperian log of Rx; =LN(Rx)

*the sheet has headers.

